# Obstructive Sleep Apnea is Associated with Peri-Lead Edema Following Deep Brain Stimulation for Parkinson’s Disease

**DOI:** 10.64898/2026.04.05.26350193

**Authors:** Evgeniya Kornilov, Uri Alkan, Ella Harari, Karam Azem, Shlomo Fireman, Eilat Kahana, Johnathan Reiner, Eilat Sapirstein, Gal Sela, Amir Glik, Shai Fein, Idit Tamir

## Abstract

**Background:** Peri-lead edema (PLE) occurs in up to 15% of Deep Brain Stimulation (DBS) cases, can cause morbidity, and its etiology remains unknown. We hypothesized that PLE represents a secondary brain injury modulated by hypoxemia, and that patients with obstructive sleep apnea (OSA) are at elevated risk.

**Methods:** We conducted a retrospective case-control study of 121 Parkinson’s disease (PD) patients undergoing DBS at a single center (2019-2024). PLE severity was quantified by CT volumetric segmentation and Hounsfield unit (HU) measures. Perioperative SpO_2_ and PaO_2_ were recorded. Polysomnography (PSG) was available in 26 patients; and the REM Sleep Behavior Disorder Screening Questionnaire (RBDSQ) was administered retrospectively.

**Results:** Symptomatic PLE occurred in 12 patients (9.9%), with onset at 3.5 (2-9) days postoperatively. PLE patients had higher body mass index (p = 0.022) and higher OSA prevalence (75% vs. 30%; p = 0.002). Perioperative SpO_2_ was lower in the PLE group in both the operating room and post-anesthesia care unit (PACU; p < 0.05); PaO_2_ was lower in the PACU (p = 0.037). In the PSG subgroup, REM Sleep Behavior Disorder (RBD) incidence was lower in PLE patients (20% vs. 60%; unadjusted p = 0.048), and PLE severity correlated significantly with sleep-related hypoxemia and respiratory indices. RBDSQ scores were positively associated with edema density (normalized HU: rho = 0.86, p = 0.024).

**Conclusions:** OSA and perioperative hypoxemia are associated with symptomatic PLE following DBS, while RBD appears protective. Preoperative sleep evaluation and optimized perioperative airway management warrant prospective investigation as PLE prevention strategies.

## Introduction

Deep Brain Stimulation (DBS) surgery is an effective and safe advanced treatment option for Parkinson’s disease (PD)^1^. For more than a decade now, reports have been published regarding a possible complication of symptomatic brain edema around the DBS implanted electrodes, often resulting in transient cognitive decline^2,3^. The so-called peri-lead edema (PLE) is often unilateral or asymmetric, and delayed in course, appearing a few days after surgery and lasts even a few weeks^4^. It is most often a self-limiting process that might partially respond to steroids treatment^5^. It occurs in at least 15% of DBS cases, but only 6% are symptomatic^2^.

Despite numerous investigations, the etiology and risk factors for PLE remain unknown, and no prevention or monitoring protocol exists. Surgical factors such as the number of microelectrodes passes as well as disease related factors have been largely excluded, and the mechanism remains obscure^4^. While MRI and animal histology demonstrate microbleeds and gliosis around implanted electrodes, routine CT rarely reveals overt pathology, and why only a subset of patients develop PLE is unexplained^6,7^.

In neurosurgery, edema in the context of traumatic brain injury (TBI) is a well described phenomenon^8,9^. It is reported to include both intra-cellular (early) and vasogenic (delayed) components, and is usually peaking around 3-7 days after the primary insult^9^. Secondary inflammatory messengers are known to mediate it. Cerebral perfusion pressure (CPP) and brain tissue oxygenation (PbtO_2_) are key determinants of secondary injury severity in TBI and are closely monitored in this setting^10^. Nevertheless, steroids to minimize brain edema are not recommended in this setting anymore, as they worsen overall prognosis due to possible systemic complications^11,12^.

We hypothesized that post-DBS PLE represents a secondary brain injury due to the minor penetrating trauma of lead insertion, modulated by systemic factors like oxygen saturation, and possibly the patient’s preoperative inflammatory status. Therefore, patients prone to hypoxemia and chronic inflammation may be at risk of developing more severe - and thus symptomatic - brain edema. PD patients possess elevated rate of obstructive sleep apnea (OSA) and sleep disturbances such as REM sleep behavior disorder (RBD), predisposing them to episodic desaturation and chronic low-grade inflammation^13–17^. Sedation during surgery may further amplify apneic tendency, aggravating hypoxemia and the inflammatory response^18^. This study investigates this hypothesis in a retrospective case-control methodology.

## Methods

### Study design, setting, and ethics

This study was conducted at Beilinson Campus of the Rabin medical center, Petach Tikva, Israel between February 2019 and January 2024 and was designed as a single center retrospective case-control trial. The study protocol was approved by the Institutional Ethics Committee (study no. 0004-20 RMC). Reporting followed the STROBE guidelines for observational research^19^.

### Study population

All consecutive patients undergoing DBS surgery targeting the subthalamic nucleus (STN) for PD during the study period were included. Patients who underwent DBS for other indications or whose procedure was performed entirely under general anesthesia were excluded. Cases were defined as patients who developed symptomatic PLE following surgery. Controls were all remaining eligible patients who did not develop symptomatic PLE during the study period.

### DBS Procedure

DBS surgery targeting the STN, was performed as previously described^20^. Briefly, the surgical procedure consisted of three stages: (1) skin incision and burr hole preparation; (2) microelectrode recordings and therapeutic window assessment; and (3) lead implantation and skin closure. Anesthesia was managed according to American Society of Anesthesiologists standard with monitored anesthesia care following the institutional protocol. Specifically, propofol sedation was used during the first and third stages, whereas during the second stage patients were either kept awake or administered an anxiolytic dose of ketamine^20^. Implantation of the internal pulse generator was performed subsequently under general anesthesia.

### Outcomes

Primary: The relationship between OSA/perioperative hypoxemia and development of symptomatic PLE following DBS surgery in PD patients. Secondary: The relationship between sleep-related hypoxemia indices and PLE severity.

### Data collection

#### Clinical and demographic data

Patient characteristics were retrospectively extracted from medical records. Collected variables included age, sex, body mass index (BMI), disease duration, Movement Disorder Society-Unified Parkinson’s Disease Rating Scale (MDS-UPDRS) scores, comorbidities, length of hospital stay, and readmissions within three months postoperatively.

#### Perioperative physiological parameters

Perioperative physiological data were extracted from the MetaVision system (iMDsoft, Tel-Aviv, Israel) and included intraoperative parameters (from the operating room, OR) and immediate postoperative parameters (from the post-anesthesia care unit, PACU). Variables included intraoperative drugs (propofol, ketamine, dexamethasone, nicardipine, phenylephrine), hemodynamics measured as mean arterial pressure (MAP), oxygenation parameters (SpO_2_) and arterial blood gases. Continuous data were automatically recorded at one-minute intervals. To account for differences in surgery duration, anesthesia drug doses were also expressed as dose rate (mg/min), calculated by dividing total dose by total surgery duration.

#### Polysomnography

Overnight polysomnography (PSG) was reviewed for patients within the study cohort. Pre-operative PSG data were available for 10 of 12 patients with symptomatic PLE and for 16 patients without PLE. In the non-PLE group, PSG referral was based on subjective sleep complaints rather than systematic screening of the full cohort. Accordingly, PSG-based comparisons were considered secondary exploratory subgroup analyses, and the non-PLE PSG subgroup should not be regarded as a random sample of all controls.

Subjective sleepiness was assessed using the Stanford Sleepiness Scale (SSS) or the Epworth Sleepiness Scale (ESS), according to data availability. Objective sleep measures included total sleep time (TST), sleep efficiency (SE), wake after sleep onset (WASO), and sleep latency (SL). The proportions of slow-wave sleep (stage N3) and rapid eye movement (REM) sleep were calculated as percentages of TST.

Oxygenation measures derived from PSG included time spent below 90% oxygen saturation (TB-90), as well as minimum and maximum oxygen saturation values recorded overnight. Respiratory variables included the respiratory disturbance index (RDI), defined as the number of apneas, hypopneas, and respiratory effort-related arousals per hour of sleep, and apnea-hypopnea indices during REM and NREM sleep (REM-AHI and NREM-AHI, respectively). REM-related OSA was defined as REM-AHI at least twice NREM-AHI, and positional OSA was defined as supine AHI at least twice non-supine AHI.

Periodic limb movements (PLM) were quantified as repetitive leg movements per hour of sleep. RBD was identified by the presence of REM sleep without atonia on electromyography together with abnormal motor behaviors observed on video-PSG.

#### Sleep questionnaires

To further assess sleep-related symptoms, three questionnaires were retrospectively administered: STOP-BANG, the REM Sleep Behavior Disorder Screening Questionnaire (RBDSQ), and the Pittsburgh Sleep Quality Index (PSQI). Nine of the 12 patients with symptomatic PLE completed the questionnaires. For comparison, 18 patients were randomly selected from the non-PLE group. Because these questionnaires were administered after the study period, these data were treated as exploratory retrospective patient-reported measures of habitual preoperative sleep and sleep-related symptoms.

#### Outcome definitions and measures

Symptomatic PLE was defined as the presence of new neurological symptoms after DBS surgery, accompanied by radiological evidence of edema surrounding the electrode on non-contrast head CT. PLE severity was quantified using clinical and radiological markers. First, the postoperative day of symptom onset was recorded (PLE POD) as a clinical marker of PLE. In most cases, non-contrast head CT was performed on the same day and used for edema quantification.

Second, edema volume (PLE volume) was calculated using iPlan software (BrainLab, version 3.0.6). Manual segmentation of the edematous region was performed on each CT slice in three dimensions to generate a volumetric model, following the approach commonly used for stereotactic radiosurgery tumor delineation.

Third, edema density was assessed using Hounsfield units (HU of PLE). Three markers were placed in the most edematous area (most hypodense), excluding the DBS lead and imaging artifacts, and averaged. Similarly, three markers were placed in unaffected white matter (either contralateral or in non-edematous ipsilateral parietal regions) and averaged (HU of normal brain). Normalized HU was calculated as the ratio of mean HU in the edematous region divided by mean HU in unaffected brain, to normalize for inter-patient variability due to technical differences between CT scans.

#### Data analysis

For continuous perioperative data, outliers and technical artifacts were removed, and missing values were reconstructed using linear interpolation. Oxygenation indices were then derived for each patient, including average SpO_2_, minimal SpO_2_, area under the curve (AUC), variability [median absolute deviation (MAD)], and cumulative time spent below 93% saturation (TB-93, minutes).

Comparisons between patients with and without PLE were performed across demographics and perioperative physiological data, polysomnography (PSG) parameters, and sleep questionnaire scores. Correlation analyses were conducted between PLE severity measures (onset, HU in PLE, normalized HU, and PLE volume; HU in normal brain served as control) and three sets of variables: perioperative oxygenation disturbances (minimal SpO_2_ and TB-93, both measured in OR and PACU), PSG oxygenation indices (TB-90, minimal SpO_2_, RDI, REM-AHI, and NREM-AHI), and RBDSQ scores.

#### Statistical analysis

All analysis was performed using custom MATLAB (R2024b, The MathWorks, Natick, MA) scripts. Given the limited sample size of patients with PLE, all analyses were performed using non-parametric methods. Data were presented as median (interquartile range) for continuous variables, and count (percentage) for categorical variables. Group comparisons were conducted using the Mann-Whitney U test for continuous data and the χ^2^ test for categorical data. Correlations were assessed using Spearman’s rank correlation test. Multiple comparisons corrections were performed with the Benjamini-Hochberg procedure where appropriate. Statistical significance was defined as p ≤ 0.05 (two-tailed).

## Results

A total of 121 PD patients who underwent DBS surgery were included (Figure 1). Symptomatic PLE developed in 12 patients (9.9%), with onset at a median of 3.5 (2-9) days postoperatively. Symptoms ranged from mild cognitive decline and dysarthria to hemiparesis, delirium, seizures, and decreased consciousness; two patients required intubation. All symptoms resolved within 3 months with oral dexamethasone and supportive care, with no surgical site infections or intracerebral bleeding. Representative CT imaging and severity measurements are shown in Figure 2. The edema volume was 37.75 (21.85-48.95) cm^3^. The HU value within the PLE region was 9.17 (5.5-12.67), compared with 33.17 (32-34.5) in normal brain tissue. The normalized HU was 0.27 (0.14-0.41). Baseline and perioperative patients’ parameters are shown in Table 1.

**Figure 1.**
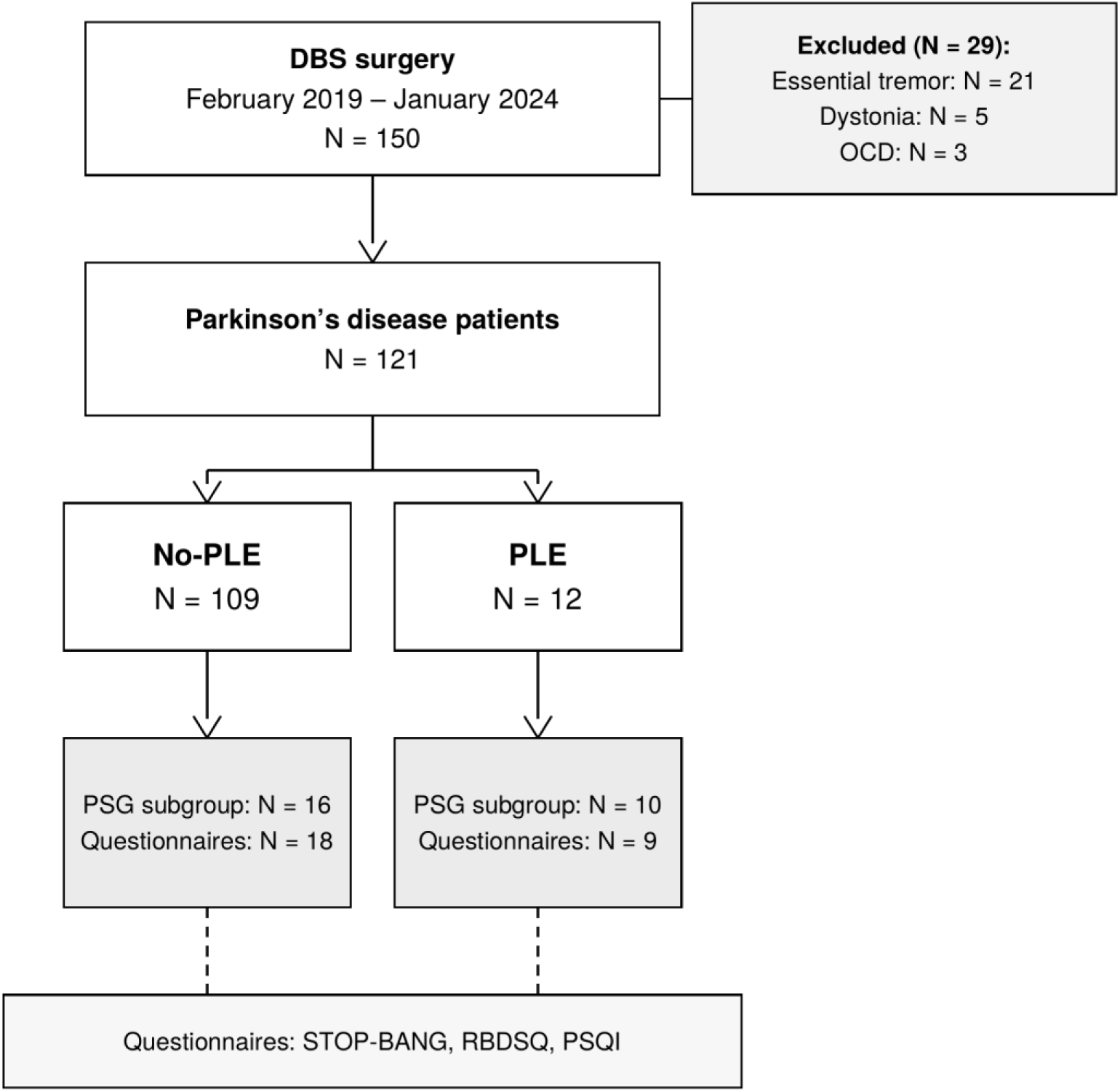
Patient selection flowchart. DBS, deep brain stimulation; PLE, peri-lead edema; PSG, polysomnography; STOP-BANG, STOP-BANG sleep apnea screening questionnaire; RBDSQ, REM Sleep Behavior Disorder Screening Questionnaire; PSQI, Pittsburgh Sleep Quality Index.

**Figure 2.**
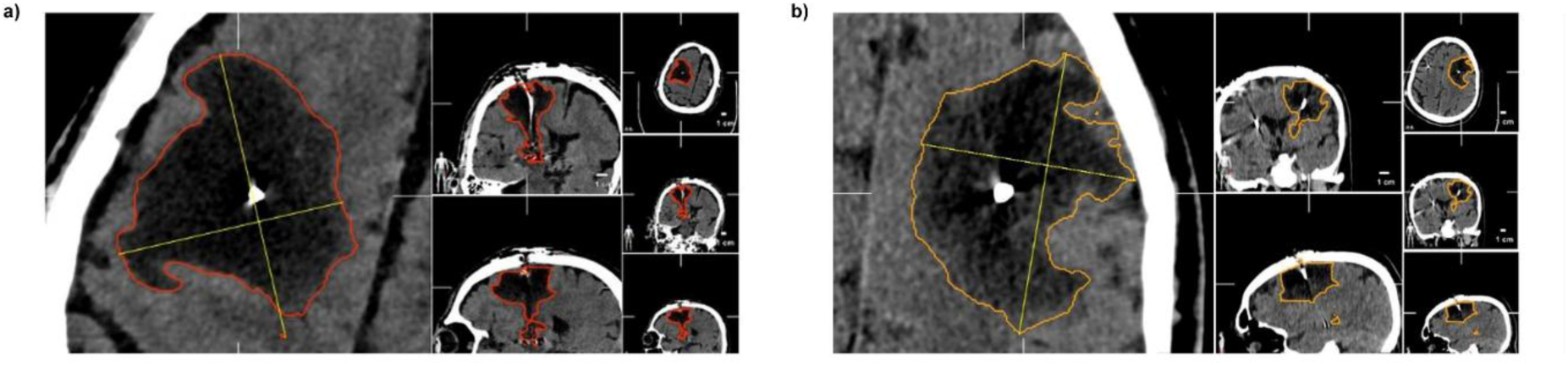
Representative CT imaging of peri-lead edema following DBS surgery. (a) Patient with edema volume of 55.2 cm³ and mean HU of 6.3. (b) Patient with edema volume of 77.0 cm³ and mean HU of 4.7. Axial (left) and multiplanar (right) views. PLE, peri-lead edema; HU, Hounsfield units; DBS, deep brain stimulation.

**Table 1.**
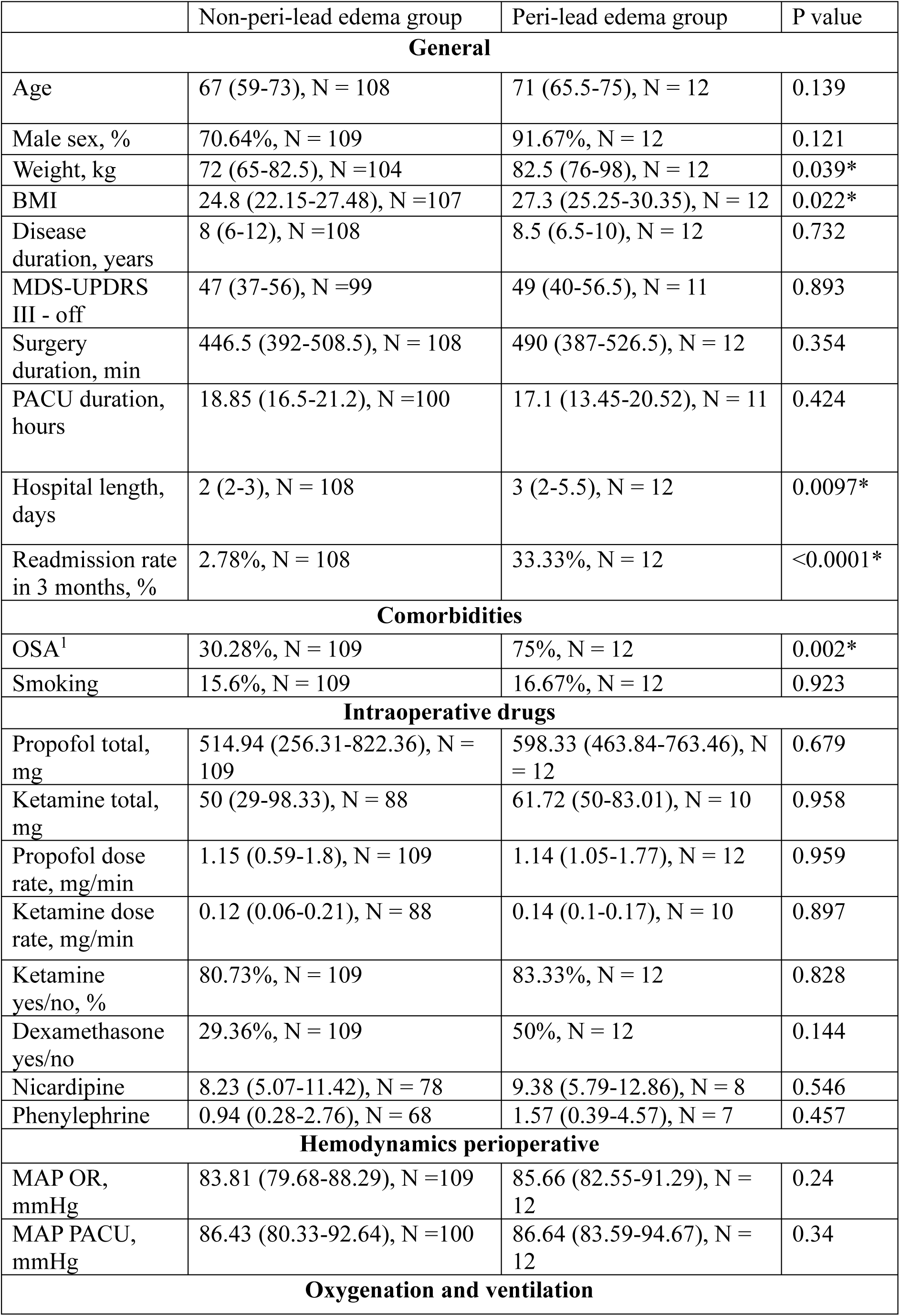
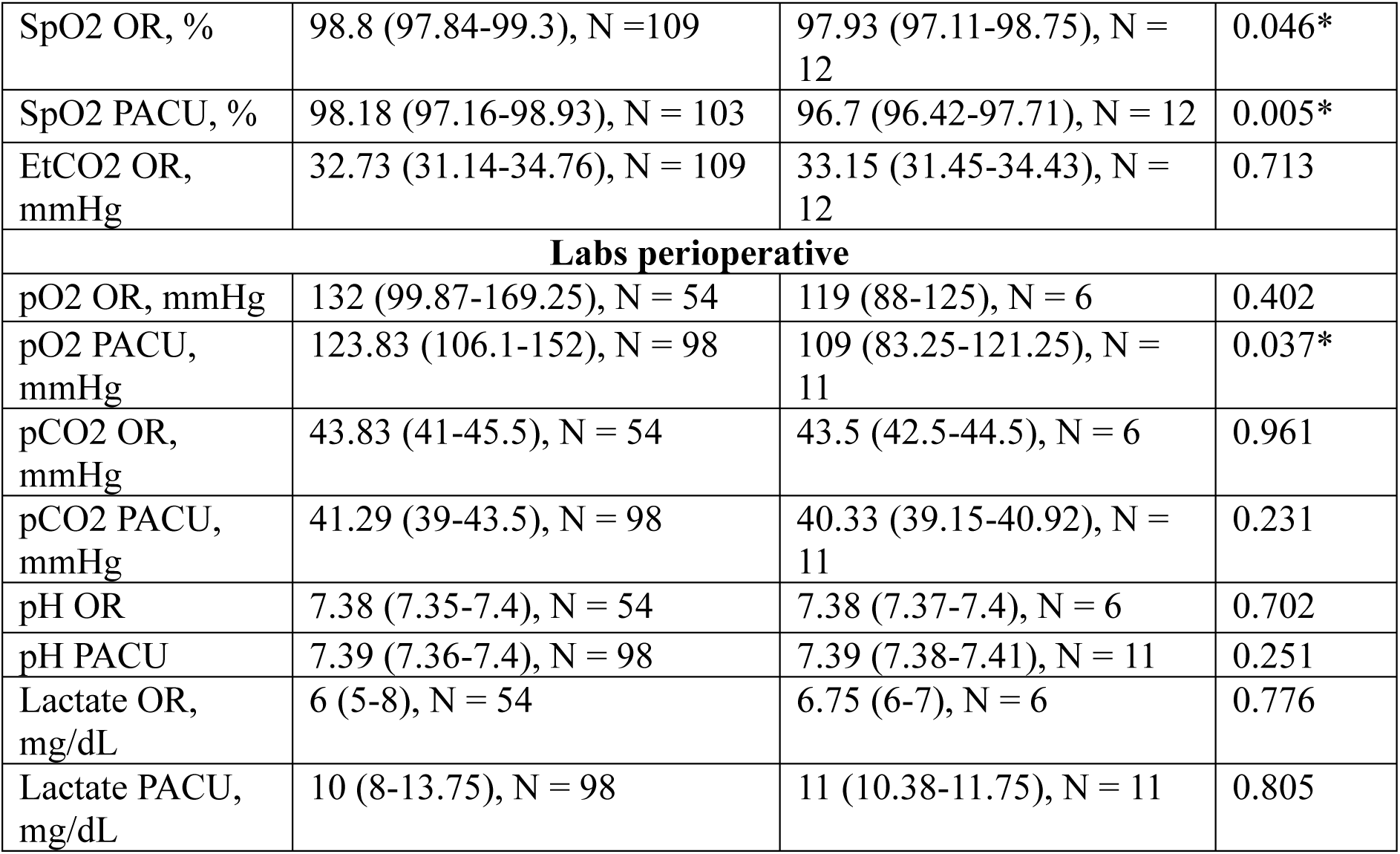
Perioperative patient characteristics in PLE and No-PLE groups. Data are presented as median (IQR) or N (%). Significance assessed with Mann-Whitney U test for continuous variables and Chi-square test for categorical variables. Significant differences are marked with *. ¹OSA as recorded in medical records prior to surgery. PLE, peri-lead edema; MDS-UPDRS III, Movement Disorder Society Unified Parkinson’s Disease Rating Scale, part III; PACU, post-anesthesia care unit; OSA, obstructive sleep apnea; MAP, mean arterial pressure; SpO_2_, peripheral oxygen saturation; EtCO_2_, end-tidal carbon dioxide; pO_2_, arterial partial pressure of oxygen; pCO_2_, arterial partial pressure of carbon dioxide; OR, operating room.

### Patients with PLE had higher incidence of OSA and lower peri-operative blood oxygenation

PLE patients had higher BMI and OSA was more than twice as common in the PLE group (75% vs 30.3%; p = 0.002). OSA is associated with recurrent desaturation episodes during sleep. Therefore, we examined whether oxygenation disturbances contributed to PLE by analyzing perioperative blood oxygenation parameters. Average, minimal SpO_2_ and AUC were lower, while SpO_2_ variability and time below 93%^21^ were higher in the PLE group in both OR and PACU (Table 1, Figure 3). PaO_2_ was additionally lower in the PACU (p = 0.037) but not intraoperatively. In PACU but not in OR, perioperative desaturation showed a trend toward association with greater PLE severity (Supplementary Figure 1).

**Figure 3.**
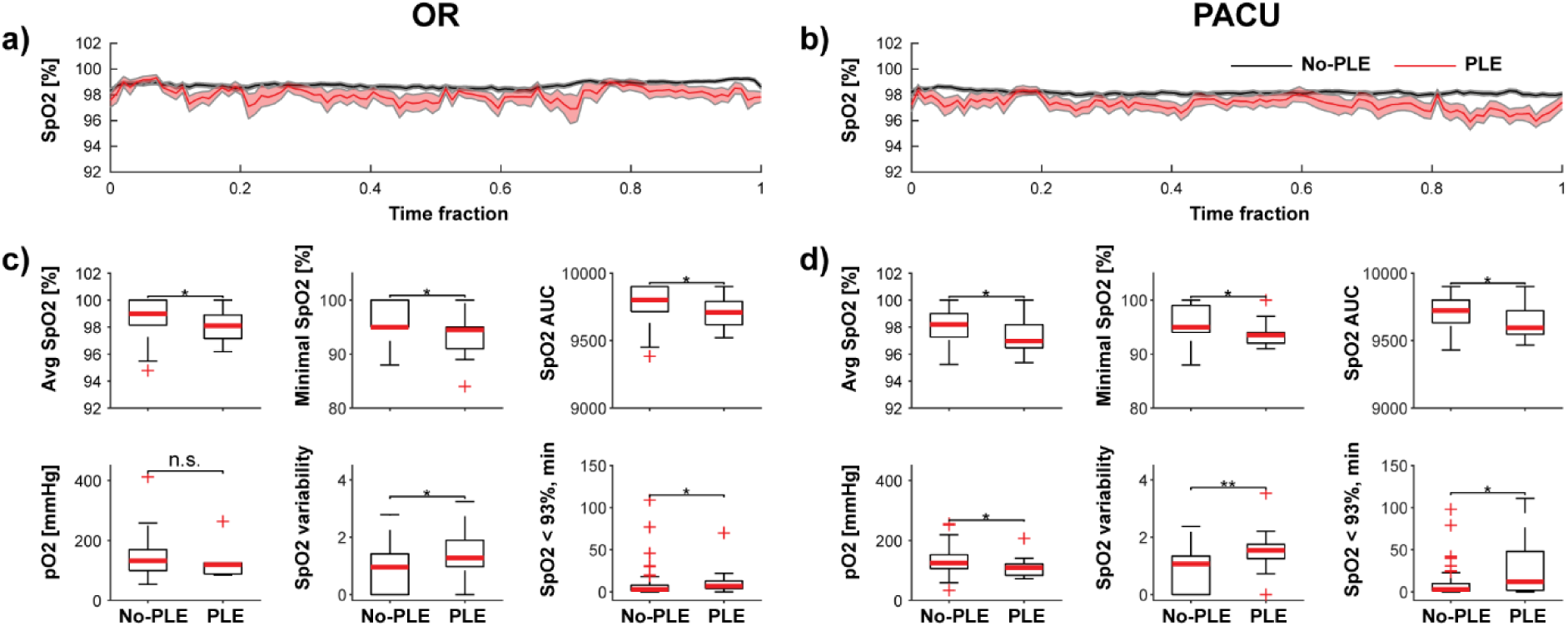
Patients with symptomatic PLE demonstrated lower perioperative blood oxygenation compared to non-PLE patients. No-PLE group is indicated in black and PLE group is indicated in red. (a, b) Mean SpO_2_ time course during the operating room (OR) and post-anesthesia care unit (PACU) periods, respectively. Shaded areas represent ± SEM. Time is expressed as a normalized fraction of total monitoring duration. (c) From left to right: average SpO_2_, %; minimal SpO_2_, %; SpO_2_ AUC; pO_2_, mmHg; SpO_2_ variability (MAD); cumulative time spent below 93% oxygen saturation, min; all measured during the OR period. (d) Same indices measured during the PACU period. Each boxplot displays the median (red line), interquartile range (box), min-max range excluding outliers (whiskers), and individual outliers (red crosses). Significance - Mann-Whitney test: n.s., p > 0.05; *, p ≤ 0.05; **, p ≤ 0.01. PLE, peri-lead edema; SpO_2_, peripheral oxygen saturation; AUC, area under the curve; pO_2_, arterial partial pressure of oxygen; OR, operating room; PACU, post-anesthesia care unit.

### Severity of PLE correlated with sleep disturbances and desaturation during sleep

PSG data were available in 26 patients (10 PLE, 16 non-PLE; Supplementary Table 1). There were trends toward higher TST, sleep efficiency and TB90 in the PLE group, showing that the PLE group slept more and had more desaturation than the non-PLE group. It is important to note that OSA patients frequently require longer total sleep time to compensate for the fragmented, poor-quality sleep caused by repeated nighttime awakenings and arousals ^22^, although some studies also report shorter overall sleep time in these patients ^23^.

PLE severity - measured by symptom onset, HU, normalized HU, and edema volume - correlated significantly with all major PSG oxygenation and respiratory indices, including TB-90, minimal SpO_2_, RDI, REM-AHI, and Non-REM-AHI (Figure 4a-d). Confirming specificity of the findings, no associations were observed between PSG parameters and HU in unaffected brain tissue (Figure 4e).

**Figure 4.**
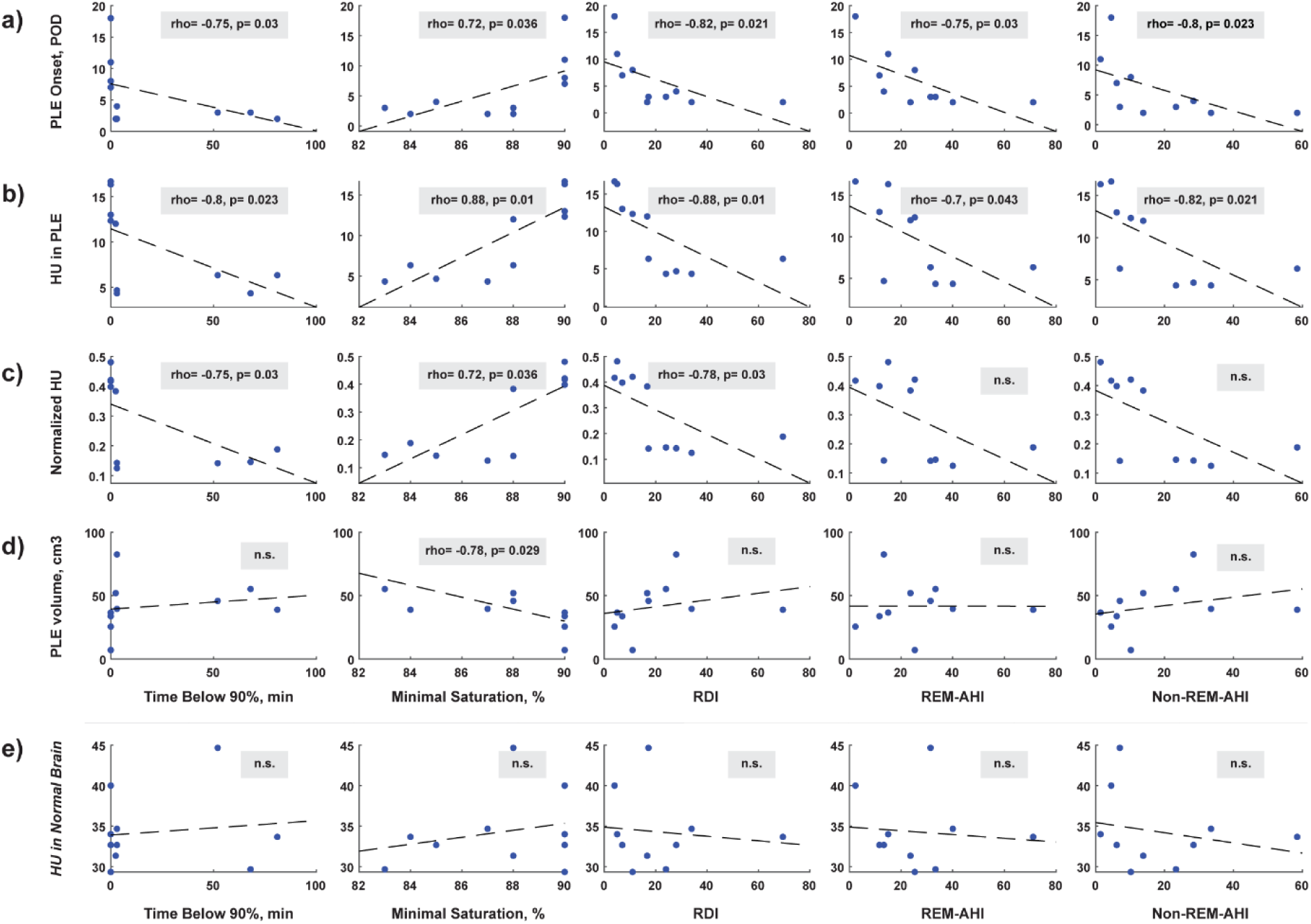
PLE severity correlated significantly with sleep-related oxygenation and respiratory indices measured by polysomnography. (a-e) Spearman correlations (rho) and corresponding p values (after post-hoc Benjamini-Hochberg procedure) between PSG-derived indices (x-axes) and four PLE severity measures (y-axes) and one control: (a) PLE onset in postoperative days; (b) Hounsfield units within the edematous region; (c) normalized HU; (d) PLE volume in cm³. (e) HU in unaffected brain. Dashed line indicates first-degree polynomial fit. n.s., p > 0.05. PLE, peri-lead edema; POD, postoperative day; HU, Hounsfield units; PSG, polysomnography; RDI, respiratory disturbance index; REM-AHI, rapid eye movement apnea-hypopnea index; Non-REM-AHI, non-rapid eye movement apnea-hypopnea index.

### Higher RBDSQ scores were associated with less severe PLE

RBD diagnosis was less frequent in PLE patients on PSG (20% vs. 60%, unadjusted p = 0.048; Figure 5f, Supplementary Table 1). Within the PLE group, higher RBDSQ scores were associated with less severe PLE: rho = 0.77 (p = 0.049) for HU and rho = 0.86 (p = 0.024) for normalized HU (Figure 5b-c), with no association observed for HU in normal brain tissue (Figure 5e). Additionally, across the full PSG cohort, REM-AHI exceeded NREM-AHI in 80.7% of patients (21/26), and was more than twice NREM-AHI in 46.2% (11/26; p < 0.002).

**Figure 5.**
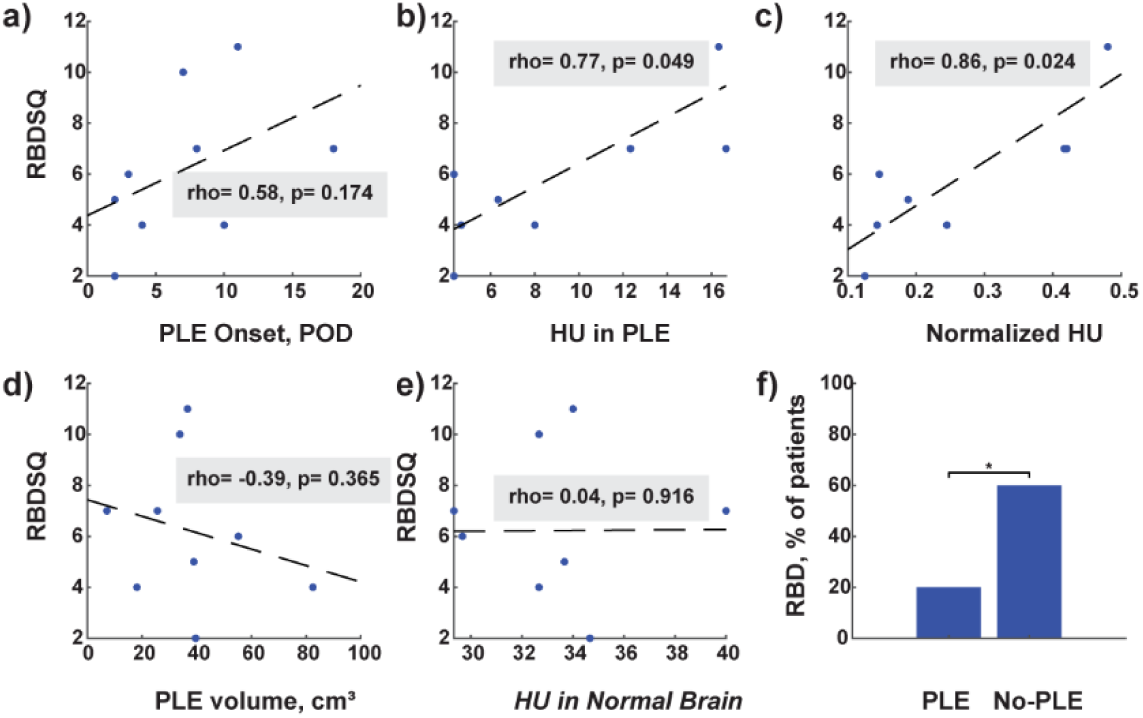
Higher RBDSQ scores were associated with less severe PLE, and RBD diagnosis was less frequent in the PLE group. (a-c) Spearman correlations (rho) and corresponding p values (after post-hoc Benjamini-Hochberg procedure) between RBDSQ score (y-axis) and PLE severity measures: (a) PLE onset in postoperative days; (b) Hounsfield units within the edematous region; (c) normalized HU. (d) PLE volume in cm³. (e) HU in unaffected brain tissue. (f) Percentage of patients with RBD diagnosis in PLE and No-PLE groups. Dashed line indicates first-degree polynomial fit. Significance - Chi-square test. *, p ≤ 0.05. PLE, peri-lead edema; POD, postoperative day; HU, Hounsfield units; RBDSQ, REM Sleep Behavior Disorder Screening Questionnaire; RBD, REM sleep behavior disorder.

## Discussion

This study provides the first evidence linking sleep-disordered breathing and perioperative hypoxemia to the development of PLE following deep brain stimulation. Patients who developed symptomatic PLE had markedly higher rates of OSA, lower perioperative oxygen saturation, and—in those with available polysomnography—strong correlations between sleep-related hypoxemia indices and edema severity. These findings suggest that PLE may represent a form of secondary brain injury, with severity modulated by the patient’s underlying tendency toward hypoxemia.

### Characteristics of post-DBS PLE

The presented results are similar to the ones previously reported in the literature^4^. In most studies, the percentage of symptomatic edema after DBS ranged from 3 to 33% and the non-symptomatic edema was much higher, up to 88-100% of patients^2,24^. Almost all patients reported by now did not develop symptoms immediately after surgery but rather at a later stage, often after home discharge^2,24^. All these cases were thought to be a result of non-infectious inflammation, like post-traumatic brain edema. Infection-related edema (i.e., cerebritis) is not self-limited and does not resolve with steroids treatment alone over time^25^. DBS is a type of penetrating trauma resulting in microbleeds and cell death along the trajectory, albeit minimal, and inducing peri-lead gliosis over time, as previously reported^26,27^. Also, the common “insertion effect” of transient symptomatic improvement after surgery is referred to local brain damage around the target with microbleeds and edema, mimicking ablation due to local disruption of neural integrity and activity^3,28,29^. Post-DBS PLE was described either in superficial subcortical areas or deep areas like the thalamus and the midbrain^24,30^. The factors determining the location of edema along the trajectory are not yet clear. Nevertheless, we found most cases of PLE to locate subcortically and disseminate deeper to the thalamus and midbrain only in more severe cases. Also, the timing of symptoms seemed to be related to the severity of edema with more severe cases presenting earlier after surgery.

### Steroids treatment in PLE

It is common to treat PLE with steroids to shorten symptom duration ^5^. We treated all symptomatic patients with steroids but did not study the effect of treatment, besides standard clinical and radiological follow up with a CT scan to ensure resolution. In neurosurgery, it is fairly common to use steroids (dexamethasone) for tumor related edema, as edema is thought to be vasogenic in nature^31^. In traumatic brain injury (TBI), steroids are considered harmful and result in worse prognosis, albeit complications are reported to be systemic and not necessarily neuronal^32^. Nevertheless, the edema of TBI originally was reported to be mainly cytotoxic and later on of a mixed type^33^. In PLE, the positive effect of steroids treatment suggests a significant component of vasogenic edema. Still, the remaining (transient) symptomatology despite treatment may be due to an additional cellular component. Our findings of apnea-related desaturation as a mechanism for the development of post DBS PLE correlates with previous findings of pro-inflammatory state in OSA patients with increased C-Reactive Protein and other markers of inflammation^34–36^. As inflammatory markers are reduced with continuous positive airway pressure (CPAP) treatment in OSA patients, we may suggest novel treatment options for PLE like the use of CPAP and possibly also prevention of apneas during surgery with close monitoring of EtCO_2_ and the use of nasal airway or general anesthesia in relevant cases. Also, conducting polysomnography before DBS to diagnose possible RBD or OSA can help identify potential patients at risk of developing post-DBS PLE. Finally, prophylactic treatment for these patients with steroids starting intra-operatively or maybe even pre-operatively may help minimize the developing edema and therefore reduce related symptoms. Another possible future option to investigate may be the use of GLP-1 agonists to reduce the OSA-related inflammatory response pre-operatively^37^.

### OSA and RBD as key factors in symptomatic peri-lead brain edema

The occurrence of OSA was twice as high in the PLE group compared to the non-PLE group. The severity of PLE was significantly correlated with the degree of sleep-related hypoxemia, as quantified by polysomnography. Patients with more severe sleep disorders had earlier PLE onset and more severe symptoms. These findings are novel and support an association between OSA and PLE consistent with a causative role, though prospective studies will be required to establish directionality.

Patients with PLE were also older and had a higher weight and BMI at the time of surgery compared with the non-PLE group. Since BMI is a well-established independent risk factor for OSA, it is possible that the association between OSA and PLE is partly mediated by or confounded by obesity. The independent contributions of BMI, OSA, and sleep-related hypoxemia to PLE risk could not be disentangled in this retrospective study, and multivariate modelling in a larger prospective cohort will be needed to resolve this.

Interestingly, while perioperative SpO_2_ was significantly lower at the group level in PLE patients both in the OR and the PACU, individual desaturation indices during these periods did not correlate significantly with PLE severity. This discrepancy is mechanistically informative: brief intraoperative hypoxemia may be insufficient on its own to drive edema severity, whereas the chronic, cumulative hypoxic burden imposed by years of sleep-disordered breathing may represent the primary modulating factor. This interpretation is consistent with the much stronger correlations observed between PSG-derived hypoxemia indices and PLE severity, and aligns with TBI literature showing that brain tissue oxygen monitoring over time - rather than single point-in-time measurements - is the key determinant of secondary injury magnitude.

In contrast to OSA, the incidence of RBD in the PLE group was lower than in the non-PLE group, although this difference did not reach significance after correction for multiple comparisons. Similarly, within the PLE group, higher RBDSQ scores were correlated with less severe edema in the small retrospective questionnaire sample. Taken together, these observations raise the exploratory hypothesis that RBD may be associated with lower PLE severity - a possibility that requires prospective confirmation in a larger cohort.

The higher incidence of OSA and RBD in PD was previously reported^14,16^. Also, age and high BMI are well known risk factors for OSA^38^. OSA in PD patients is usually related more to dopamine dose taken and upper airway muscle involvement in the disease^39,40^ rather than the common metabolic risk factors in the general population, even though metabolic syndrome is a risk factor to develop PD^41^, and also may be related to decreased dopaminergic availability at the caudate nucleus in these patients^40^. Yet the mechanism of OSA in the PD population is not completely clear.

The relationship between OSA and RBD is particularly interesting, as there is no strict evidence regarding the occurrence of OSA in PD in relation to sleep stage^42,43^. We offer the following post-hoc mechanistic interpretation, as the protective role of RBD was not a pre-specified hypothesis of this study. It is possible that PD patients have predominantly REM-related OSA, with a higher apnea burden during REM than NREM sleep — a pattern consistent with the REM-AHI predominance observed in 80.7% of our PSG cohort. In this context, the protective effect of RBD against PLE may result from frequent arousals or prevention of upper airway muscle collapse during REM sleep, limiting the duration of individual apnea events and thereby attenuating cumulative desaturation. Several studies have previously suggested a protective effect of REM sleep without atonia against OSA in PD patients, attributed to repeated awakenings during REM sleep^43^. More research aimed at this question is warranted to better understand this issue.

### Study Limitations

This study was conducted retrospectively. Therefore, no randomization or blinding were used. Also, due to the nature of this study we did not look carefully at EtCO_2_ and PaO_2_ during surgery. We did not test preventive measures or treatment regimens to reduce the occurrence of PLE. We did not send all patients to PSG study pre-operatively as this is not a recommended routine in DBS. Therefore, a few of the PLE group patients completed PSG after surgery. This potentially may affect the study results. In addition, as most of our patients undergo DBS under sedation (propofol-ketamine) and due to the retrospective nature of the study, we did not study the effect of sedation/type of anesthesia on PLE rate. The patients without symptomatic PLE that underwent PSG were the ones that complained about sleep problems when asked. Therefore, the occurrence of OSA and RBD in the general population of PD patients undergoing DBS is probably lower, and the occurrence in the general PD population (not only the ones referred to DBS) is maybe even lower than that. OSA diagnosis in the primary cohort was based on pre-existing medical records rather than standardized pre-operative testing, introducing potential ascertainment bias, as patients with higher BMI may have been more likely to have received a prior OSA diagnosis. Finally, sleep questionnaires were administered retrospectively after the study period; although items address habitual sleep prior to surgery, recall bias cannot be excluded, particularly in patients who experienced significant postoperative neurological symptoms.

## Conclusions

OSA and perioperative hypoxemia are associated with symptomatic PLE following DBS, while RBD appears to be protective. These findings suggest that post-DBS PLE represents a form of secondary brain injury modulated by chronic sleep-related hypoxemia. Pre-operative polysomnography and optimized airway management perioperatively warrant prospective evaluation as PLE prevention strategies.

## Data Availability

The datasets analyzed during the current study are not publicly available due to participant privacy regulations but are available from the corresponding author on reasonable request.

## Acknowledgements

This study received no funding. We thank Dr. Iskhakova Liliya for critical reading and comments on the manuscript.

## Author contributions

E.Ko. and I.T. conceived and designed the study. E.Ko. performed data collection, statistical analysis, and wrote the manuscript. U.A. contributed to sleep laboratory data collection and interpretation of polysomnography findings. E.H. and E.S. contributed to CT volumetric segmentation and neurosurgical data collection. K.A., S.F. and E.Ka. contributed to perioperative physiological data collection and clinical anesthesia management. J.R., G.S. and A.G. contributed to neurological assessment, clinical follow-up of PLE patients, and data collection. S.Fe. and I.T. critically revised the manuscript. I.T. supervised the study. All authors read and approved the final manuscript.

## Code Availability

MATLAB code used is available from the corresponding author on reasonable request.

## References

1 Hitti, F. L. et al. Long-term outcomes following deep brain stimulation for Parkinson’s disease. J Neurosurg 132, 205–210, doi:10.3171/2018.8.JNS182081 (2020).

2 Whiting, A. C. et al. Peri-Lead Edema After Deep Brain Stimulation Surgery: A Poorly Understood but Frequent Complication. World Neurosurg, doi:10.1016/j.wneu.2018.12.092 (2018).

3 Nishiguchi, Y. et al. Relationship of brain edema after deep brain stimulation surgery with motor and cognitive function. Heliyon 8, e08900, doi:10.1016/j.heliyon.2022.e08900 (2022).

4 Giordano, M. et al. Incidence and management of idiopathic peri-lead edema (IPLE) following deep brain stimulation (DBS) surgery: Case series and review of the literature. Clin Neurol Neurosurg 234, 108009, doi:10.1016/j.clineuro.2023.108009 (2023).

5 Wu, B. et al. Postoperative use of steroids for peri-electrode edema after deep brain stimulation surgery: A retrospective cohort study. CNS Neurosci Ther 30, e14470, doi:10.1111/cns.14470 (2024).

6 Vedam-Mai, V. et al. Deep Brain Stimulation associated gliosis: A post-mortem study. Parkinsonism Relat Disord 54, 51–55, doi:10.1016/j.parkreldis.2018.04.009 (2018).

7 Vivanco-Suarez, J. et al. Neurohistopathological findings of the brain parenchyma after long-term deep brain stimulation: Case series and systematic literature review. Parkinsonism Relat Disord 133, 107243, doi:10.1016/j.parkreldis.2024.107243 (2025).

8 Zusman, B. E., Kochanek, P. M. & Jha, R. M. Cerebral Edema in Traumatic Brain Injury: a Historical Framework for Current Therapy. Curr Treat Options Neurol 22, doi:10.1007/s11940-020-0614-x (2020).

9 Ramirez de Noriega, F., et al. A swine model of intracellular cerebral edema - Cerebral physiology and intracranial compliance. J Clin Neurosci 58, 192–199, doi:10.1016/j.jocn.2018.10.051 (2018).

10 Shen, Y. et al. Brain tissue oxygen partial pressure monitoring and prognosis of patients with traumatic brain injury: a meta-analysis. Neurosurg Rev 47, 222, doi:10.1007/s10143-024-02439-4 (2024).

11 Prasad, G. L., Pai, A. & Pt, S. Short course of low-dose steroids for management of delayed pericontusional edema after mild traumatic brain injury - A retrospective study. Surg Neurol Int 16, 23, doi:10.25259/SNI_948_2024 (2025).

12 Edwards, P. et al. Final results of MRC CRASH, a randomised placebo-controlled trial of intravenous corticosteroid in adults with head injury-outcomes at 6 months. Lancet 365, 1957–1959, doi:10.1016/S0140-6736(05)66552-X (2005).

13 Maggi, G., Giacobbe, C., Iannotta, F., Santangelo, G. & Vitale, C. Prevalence and clinical aspects of obstructive sleep apnea in Parkinson disease: A meta-analysis. Eur J Neurol 31, e16109, doi:10.1111/ene.16109 (2024).

14 Shen, Y. et al. Obstructive sleep apnea in Parkinson’s disease: a study in 239 Chinese patients. Sleep Med 67, 237–243, doi:10.1016/j.sleep.2019.11.1251 (2020).

15 Martinez-Nunez, A. E. et al. Clinically probable RBD is an early predictor of malignant non-motor Parkinson’s disease phenotypes. NPJ Parkinsons Dis 11, 25, doi:10.1038/s41531-025-00874-8 (2025).

16 Postuma, R. B., Gagnon, J. F., Bertrand, J. A., Genier Marchand, D. & Montplaisir, J. Y. Parkinson risk in idiopathic REM sleep behavior disorder: preparing for neuroprotective trials. Neurology 84, 1104–1113, doi:10.1212/WNL.0000000000001364 (2015).

17 Kubler-Weller, D. et al. Predicting cognition after subthalamic Deep Brain Stimulation in Parkinson’s Disease. NPJ Parkinsons Dis 11, 265, doi:10.1038/s41531-025-01128-3 (2025).

18 Ceban, F. et al. Adverse events in patients with obstructive sleep apnea undergoing procedural sedation in ambulatory settings: An updated systematic review and meta-analysis. Sleep Med Rev 80, 102029, doi:10.1016/j.smrv.2024.102029 (2025).

19 von Elm, E. et al. The Strengthening the Reporting of Observational Studies in Epidemiology (STROBE) statement: guidelines for reporting observational studies. Lancet 370, 1453–1457, doi:10.1016/S0140-6736(07)61602-X (2007).

20 Kornilov, E. et al. Interleaved Propofol-Ketamine Maintains DBS Physiology and Hemodynamic Stability: A Double-Blind Randomized Controlled Trial. Mov Disord 39, 694–705, doi:10.1002/mds.29746 (2024).

21 Nordenholz, K., Ryan, J., Atwood, B. & Heard, K. Pulmonary embolism risk stratification: pulse oximetry and pulmonary embolism severity index. J Emerg Med 40, 95–102, doi:10.1016/j.jemermed.2009.06.004 (2011).

22 Oksenberg, A. & Leppanen, T. Duration of respiratory events in obstructive sleep apnea: In search of paradoxical results. Sleep Med Rev 68, 101728, doi:10.1016/j.smrv.2022.101728 (2023).

23 Risso, T. T. et al. The impact of sleep duration in obstructive sleep apnea patients. Sleep Breath 17, 837–843, doi:10.1007/s11325-012-0774-3 (2013).

24 Borellini, L. et al. Peri-lead edema after deep brain stimulation surgery for Parkinson’s disease: a prospective magnetic resonance imaging study. Eur J Neurol 26, 533–539, doi:10.1111/ene.13852 (2019).

25 Fishman, R. A. Steroids in the treatment of brain edema. N Engl J Med 306, 359–360, doi:10.1056/NEJM198202113060609 (1982).

26 Nielsen, M. S. et al. Chronic subthalamic high-frequency deep brain stimulation in Parkinson’s disease--a histopathological study. Eur J Neurol 14, 132–138, doi:10.1111/j.1468-1331.2006.01569.x (2007).

27 Moss, J., Ryder, T., Aziz, T. Z., Graeber, M. B. & Bain, P. G. Electron microscopy of tissue adherent to explanted electrodes in dystonia and Parkinson’s disease. Brain 127, 2755–2763, doi:10.1093/brain/awh292 (2004).

28 Mann, J. M. et al. Brain penetration effects of microelectrodes and DBS leads in STN or GPi. J Neurol Neurosurg Psychiatry 80, 794–797, doi:10.1136/jnnp.2008.159558 (2009).

29 Lawson McLean, A. & Nemir, J. Quantifying insertional effects in deep brain stimulation: clinical outcomes and neurophysiological mechanisms. Expert Rev Med Devices 22, 285–291, doi:10.1080/17434440.2025.2480660 (2025).

30 Johnson, C. et al. Unveiling patterns of peri-lead edema after deep brain stimulation: a retrospective review of clinical and demographic factors. Neuroradiology, doi:10.1007/s00234-025-03607-z (2025).

31 Roth, P., Happold, C. & Weller, M. Corticosteroid use in neuro-oncology: an update. Neurooncol Pract 2, 6–12, doi:10.1093/nop/npu029 (2015).

32 Alderson, P. & Roberts, I. Corticosteroids for acute traumatic brain injury. Cochrane Database Syst Rev 2005, CD000196, doi:10.1002/14651858.CD000196.pub2 (2005).

33 Lee, J. et al. Edema progression in proximity to traumatic microbleeds: evolution of cytotoxic and vasogenic edema on serial MRI. Neuroimage Rep 4, doi:10.1016/j.ynirp.2024.100199 (2024).

34 Lundetrae, R. S. et al. Severity of obstructive sleep apnea is related to C-reactive protein levels: The influence of comorbidities and self-reported sleep duration. Sleep Med 131, 106529, doi:10.1016/j.sleep.2025.106529 (2025).

35 Nadeem, R. et al. Serum inflammatory markers in obstructive sleep apnea: a meta-analysis. J Clin Sleep Med 9, 1003–1012, doi:10.5664/jcsm.3070 (2013).

36 Shamsuzzaman, A. S. et al. Elevated C-reactive protein in patients with obstructive sleep apnea. Circulation 105, 2462–2464, doi:10.1161/01.cir.0000018948.95175.03 (2002).

37 Malhotra, A. et al. Tirzepatide for the Treatment of Obstructive Sleep Apnea and Obesity. N Engl J Med 391, 1193–1205, doi:10.1056/NEJMoa2404881 (2024).

38 Messineo, L., Bakker, J. P., Cronin, J., Yee, J. & White, D. P. Obstructive sleep apnea and obesity: A review of epidemiology, pathophysiology and the effect of weight-loss treatments. Sleep Med Rev 78, 101996, doi:10.1016/j.smrv.2024.101996 (2024).

39 Yu, Q. et al. Obstructive sleep apnea in Parkinson’s disease: A prevalent, clinically relevant and treatable feature. Parkinsonism Relat Disord 115, 105790, doi:10.1016/j.parkreldis.2023.105790 (2023).

40 Oh, Y. S., Kim, J. S., Lyoo, C. H. & Kim, H. Obstructive Sleep Apnea and Striatal Dopamine Availability in Parkinson’s Disease. Mov Disord 38, 1068–1076, doi:10.1002/mds.29402 (2023).

41 Zhong, Y., Wang, T. H., Huang, L. J. & Hua, Y. S. Association between metabolic syndrome and the risk of Parkinson’s disease: a meta-analysis. BMC Neurol 24, 313, doi:10.1186/s12883-024-03820-y (2024).

42 Jung, Y. J. & Oh, E. Is REM sleep behavior disorder a friend or foe of obstructive sleep apnea? Clinical and etiological implications for neurodegeneration. J Clin Sleep Med 17, 1305–1312, doi:10.5664/jcsm.9144 (2021).

43 Jo, S., Kim, H. W., Jeon, J. Y. & Lee, S. A. Protective effects of REM sleep without atonia against obstructive sleep apnea in patients with idiopathic REM sleep behavior disorder. Sleep Med 54, 116–120, doi:10.1016/j.sleep.2018.10.032 (2019).

