## Supplementary Material for "Obstructive Sleep Apnea is Associated with Peri-Lead Edema Following Deep Brain Stimulation for Parkinson’s Disease"

| | Non-peri-lead edema group | Peri-lead edema group | P value | P value adjusted |
|---|---|---|---|---|
| **Sleep lab parameters** | | | | |
| SSS/ESS | 6 (4-9.5), n = 16 | 8.5 (4-11), n = 10 | 0.431 | 0.867 |
| TST, hours | 4 (3.18-5), n = 15 | 5.5 (5.1-5.6), n = 10 | 0.019 | 0.323 |
| Sleep Efficiency | 62 (49.25-75.25), n = 15 | 76.75 (68-84), n = 10 | 0.094 | 0.386 |
| WASO | 32 (12-83), n = 12 | 25 (6.88-83), n = 11 | 0.766 | 0.867 |
| Sleep Latency | 40 (19-96.75), n = 15 | 34.75 (18-90), n = 10 | 0.37 | 0.867 |
| TB-90, min | 0 (0-6), n = 15 | 2.7 (0-52), n = 10 | 0.113 | 0.386 |
| Maximal Saturation | 98 (97-98), n = 15 | 97.5 (97-98), n = 10 | 0.198 | 0.56 |
| Minimal Saturation | 90 (88-90), n = 15 | 88 (85-90), n = 10 | 0.859 | 0.867 |
| RDI | 15.5 (6-26.3), n = 14 | 16.95 (7-28), n = 10 | 0.648 | 0.867 |
| REM-AHI | 20.9 (13-44.4), n = 11 | 24.45 (13.3-33.3), n = 10 | 0.834 | 0.867 |
| Non-REM-AHI | 15.1 (2.15-16.4), n = 11 | 12 (6.1-28.4), n = 10 | 0.642 | 0.867 |
| N3 | 18.7 (11.4-26), n = 14 | 19.75 (8.4-22), n = 10 | 0.462 | 0.867 |
| REM | 16.8 (0-26), n = 14 | 9.6 (6-28.9), n = 10 | 0.862 | 0.867 |
| PLM | 2 (0-3), n = 13 | 4 (1-10), n = 10 | 0.075 | 0.386 |
| RBD | 60%, n = 15 | 20%, n = 10 | 0.048 | 0.386 |
| ROSA | 50%, n = 10 | 40%, n = 8 | 0.671 | 0.867 |
| POSA | 10%, n = 10 | 12.5%, n = 8 | 0.867 | 0.867 |
| **Sleep questionnaires** | | | | |
| RBDSQ | 6.5 (4-9), n = 18 | 6 (4-7.75), n = 9 | 0.812 | — |
| STOP-BANG | 4 (3-5), n = 18 | 4 (2.75-4.25), n = 9 | 0.608 | — |
| PSQI | 9 (7-10), n = 18 | 8 (4.75-10.25), n = 9 | 0.591 | — |

**Supplementary Table 1. Sleep lab parameters and sleep questionnaires comparisons**. Data presented as median (IQR) or %. SSS/ESS - Stanford Sleepiness Scale/Epworth Sleepiness Scale; TST - Total Sleep Time; WASO - Wake After Sleep Onset; TB-90 - Time below 90% oxygen saturation; RDI - Respiratory Disturbance Index; REM-AHI - Rapid Eye Movement Apnea–Hypopnea Index; Non-REM-AHI - Non–Rapid Eye Movement Apnea–Hypopnea Index; N3 - Slow-wave sleep; REM - Rapid Eye Movement Sleep; PLM - Periodic Limb Movements; RBD - REM Behavior Disorder; ROSA - REM-related Obstructive Sleep Apnea; POSA - Positional Obstructive Sleep Apnea; RBDSQ - REM Sleep Behavior Disorder Screening Questionnaire; STOP-BANG - STOP-BANG Questionnaire; PSQI -

Pittsburgh Sleep Quality Index. P value - Mann-Whitney U test for continuous variables; $\chi^2$ test for categorical variables. P value adjusted - The Benjamini–Hochberg procedure.

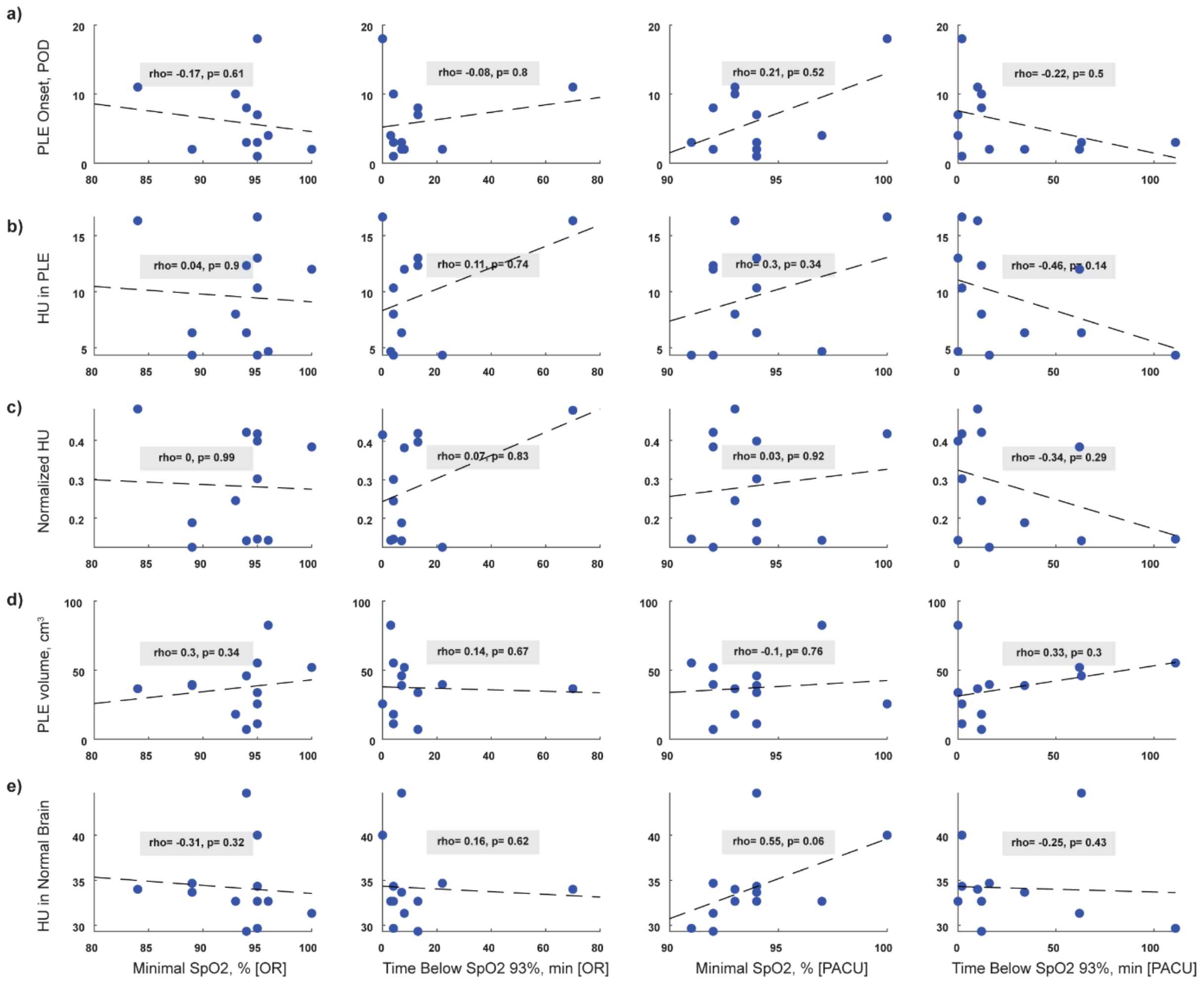


**Supplementary Figure 1. In PACU, perioperative desaturation showed a trend toward association with greater PLE severity.**

(a–d) Spearman correlations (rho) and corresponding p values (p) between perioperative $SpO_2$-derived indices (x-axes) and four PLE severity measures (y-axes): (a) PLE onset in postoperative days; (b) Hounsfield units within the edematous region; (c) normalized HU (HU in edematous region divided by HU in unaffected brain); (d) PLE volume in $cm^3$. (e) HU in unaffected brain tissue, serving as a negative control. Columns represent, from left to right: minimal $SpO_2$ during OR; cumulative time below $SpO_2$ 93% during OR; minimal $SpO_2$ during PACU; cumulative time below $SpO_2$ 93% during PACU. Dashed line indicates first-degree polynomial fit. PLE, peri-lead edema; POD, postoperative day; HU, Hounsfield units;

$SpO_2$, peripheral oxygen saturation; OR, operating room; PACU, post-anesthesia care unit. P values are uncorrected for multiple comparisons.
